# Predicting Chemotherapy Response from Staging Laparoscopy Images

**DOI:** 10.64898/2026.06.22.26356226

**Authors:** Thomas Schnelldorfer, Janil Castro, Atoussa Goldar-Najafi, Francis W. Nugent, Bipin Gaikwad

## Abstract

**Background:** For patients with metastatic gastrointestinal cancers, chemotherapy resistance is a common phenomenon that, if known in advance, would allow for individualized treatment decisions. This study aimed to test the feasibility of developing a deep learning computer vision system that uses laparoscopy images depicting peritoneal surface metastases (i.e., capturing the in-vivo optical appearance of metastases as a summary of their molecular makeup) to predict whether a patient is resistant to standard chemotherapy.

**Methods:** The retrospective observational feasibility study included 35 adult patients who underwent staging laparoscopy for non-colon gastrointestinal adenocarcinoma with biopsy-confirmed peritoneal surface metastases and who underwent chemotherapy as their only treatment modality. Chemotherapy resistance was determined based on each patient’s observed cancer-specific survival after controlling for confounders.

**Results:** Of 35 patients, 17 were assigned to the chemotherapy sensitive group and 18 to the chemotherapy resistant group. The study cohort provided 1010 laparoscopy image patches of 101 biopsy-confirmed metastases. A densely connected convolutional neural network with cross-validation provided the best results for correctly predicting chemotherapy resistance at the patient level (accuracy 0.80 (95%CI 0.63-0.92), sensitivity 0.72, specificity 0.88, AUC-ROC 0.78). Saliency maps demonstrated the system’s trustworthiness.

**Conclusion:** In this study, a prototype surgical computer vision system designed to determine chemotherapy resistance from operative images of peritoneal surface metastases demonstrated its technical feasibility. Further development and validation in a multi-institutional clinical study are pending.

## Introduction

Cancer metastases exhibit extensive variability in clonality, providing a variety of responses to specific chemotherapy regimens.^1^ Predicting patient-specific responses to a given regimen remains challenging. Clinical trials can reasonably predict what works on average in similar patient cohorts defined by limited clinical markers. However, since no patient is average and since these limited clinical markers result in fairly heterogeneous groups, there is currently no reliable method to predict chemotherapy response for individual patients.

Prior approaches to solve this challenge include the identification of molecular and clinical data biomarkers, formation of tumor organoids and xenograft models, whole-genome sequencing, and analysis of radiographic and histology images. Yet, currently, none of these approaches have been validated in larger cohorts, likely due to various reasons such as the artificial environment of the method used, only representing a limited cancer location with a specific set of clones, or sampling of a limited number of clones, the unpredictability of epigenetics, and the fact that many mutations are not vetted regarding their role within chemotherapy resistance.^2–14^ Even attempts to improve trial precision through N-of-1 trials, umbrella trials, platform trials, and biomarker guided trials have not and are not expected to reach the goal of true precision cancer care.^15,16^ Since the optical appearance of cancer tissue represents a summary of its molecular makeup, in vivo imaging of metastases may offer guidance.^17^ Standard laparoscopic imaging of peritoneal surface metastases (PSM) has the potential to capture these optical properties in their natural environment though a minimal invasive approach. Because of our recent successes with a computer vision surgical guidance system that is able to identify PSM on standard laparoscopy images,^17^ the question was raised whether a computer vision system that uses the optical appearance of PSMs depicted in its natural environment on standard laparoscopy images is able to predict resistance to standard chemotherapy regimens. The aim of this retrospective observational feasibility study, therefore, was to develop a foundational pathway of how to build a deep learning system capable of predicting chemotherapy resistance from standard laparoscopy images depicting PSMs.

## Methods

### Data Source

For this study, an established image library containing conventional cancer staging laparoscopy videos performed by a single surgeon (T.S.) at two tertiary care hospitals under the standard of care between January 2014 and October 2023 was utilized. Patients who met the following inclusion criteria were recruited from the library: 1) adult patients (≥18 year of age) with non-colon gastrointestinal adenocarcinoma, 2) received cytotoxic chemotherapy as the only cancer treatment, 3) available video recording the entire length of the staging laparoscopy performed as part of routine medical care, 4) underwent at least one intra-operative biopsy histologically confirming the presence of a PSM, 5) patient signed a consent form that includes consent for video recording. Individuals who had impaired capacity to make informed medical decisions were not included in the library. From the patient’s electronic medical record, clinical data relevant to their cancer treatment were obtained. Seventy-nine percent of histopathology slides of biopsied peritoneal surface lesions were available for re-review by a single gastrointestinal pathologist (A.G.N.). Discrepancies with the original pathology result were resolved by group consensus. The study was approved by the Lahey Clinic and Tufts Medical Center IRB.

### Preprocessing and Annotation of Data

#### Laparoscopy Images

For the development of the deep learning system, the following dataset was curated. Ten still images best representing each biopsied PSM were obtained from each laparoscopy video. These still images were selected by the greatest optical variance, depicting different angles and imaging distances within each laparoscopy video. Image patches were obtained by cropping each image to tightly include only the region of interest / biopsied PSM. These 10 image patches per each biopsied PSM were subsequently used for the development of the system.

All images were processed by a trained annotation specialist (J.C.) and supervised by an oncologic surgeon (T.S.). All laparoscopy images were stored in PNG format (24-bit RGB, lossless compression). The quality of the images was assured by only obtaining images with a clear / non-obstructed view, sufficient illumination, region of interest in-focus, and good resolution. All image patches underwent Z-score normalization for each color channel based on combined mean and standard deviation values from the entire still image sample and were subsequently resized to an 80x80 pixel format. All image patches were stored in a CSV format.

#### Determination of Chemotherapy Resistance

Patients were labeled chemotherapy sensitive or resistant based on actual versus expected cancer-specific survival within strata controlled for confounding. Cancer-specific survival was used as a surrogate for chemotherapy resistance. To avoid confounding by features that influenced survival unrelated to chemotherapy-resistance, significant predictors of cancer-specific survival within the study cohort were identified. Patients were then stratified according to any significant confounders. For each stratum, the median cancer-specific survival was calculated using Kaplan-Meier estimates. Each patient’s actual cancer-specific survival was then compared to the median cancer-specific survival within the relevant stratum. Based on this comparison, study patients were assigned to one of two groups: ‘chemotherapy sensitive’ (i.e., patient’s actual cancer-specific survival was greater than the median cancer-specific survival within its stratum) or ‘chemotherapy resistant’ (i.e., patient’s actual cancer-specific survival was less than the median cancer-specific survival within its stratum). All image patches were labeled according to the group assignment of the corresponding study patient. These labeled image patches represented the data used for training and testing of the deep neural networks.

### Model Development, Computational Environment, and Statistical Analysis

Multiple deep learning systems with various architectures were tested. Development of the deep learning systems was carried out on an HPC cluster by allocating 16 cores, 32GB memory, and an NVIDIA v100 GPU. The system’s outcome measure was accuracy, sensitivity, and specificity at a prediction threshold of 0.5 and the area under the receiver operating characteristic curve (AUC-ROC) of the system to correctly classify chemotherapy resistance. For univariate analysis, Fisher’s exact test and Student’s t-test were used. Cancer-specific survival was calculated from the start of treatment (i.e., chemotherapy) using Kaplan-Meier estimates. For group comparison of survival data, a Cox proportional hazards model was used. All hypothesis tests were 2-sided and a p-value less than 0.05 was considered significant. Statistical analyses were performed using MATLAB software version R2023a (MathWorks, Natick, MA).

## Results

### Study Demographic

The study demographics of the 35 patients who met eligibility criteria are listed in Table 1. During the study’s observation period, 29 patients died from cancer and 6 were alive at the last follow-up. The median cancer-specific survival from the start of the first chemotherapy was 17.0 months, with a 1-year cancer-specific survival of 56%. Within this cohort, 101 peritoneal surface metastases were biopsied, involving the parietal peritoneum (n=86), liver surface (n=8), and other visceral surface sites (n=7). The dataset, therefore, provided 1010 image patches (10 per biopsied PSM from different angles and distances) that were utilized for training and testing of the system. The mean peritoneal cancer index was 6 +/- 5.

**Table 1:** Study demographics.

|  | Chemotherapy<br>Sensitive Group<br>(n=17) | Chemotherapy<br>Resistant Group<br>(n=18) | p-value |
| --- | --- | --- | --- |
| **Patient Factors** |  |  |  |
| Sex | 16 males | 13 males | 0.177 * |
| Mean age | 63 +/- 9 years | 63 +/- 10 years | 0.972 ** |
| Preoperative ECOG performance score |  |  | 0.668 * |
| ECOG 0 | 11 patients | 14 patients |  |
| ECOG 1 | 4 patients | 2 patients |  |
| ECOG 2 | 2 patients | 2 patients |  |
| Mean body mass index | 27 +/- 6 kg/m<sup>2</sup> | 26 +/- 4 kg/m<sup>2</sup> | 0.447 ** |
| **Disease Factors** |  |  |  |
| Cancer type |  |  | 0.642 * |
| Gastric adenocarcinoma | 8 patients | 9 patients |  |
| Pancreatic adenocarcinoma | 6 patients | 8 patients |  |
| Other non-colon gastrointestinal<br>adenocarcinoma | 3 patients | 1 patient |  |
| Location of biopsied PSM |  |  | 0.263 * |
| Parietal peritoneum | 45 metastases | 41 metastases |  |
| Liver and other visceral surfaces | 5 metastases | 10 metastases |  |
| Mean peritoneal cancer index | 7 +/- 6 | 5 +/- 4 | 0.335 ** |
| Degree of malignant ascites |  |  | 0.443 * |
| None | 11 patients | 14 patients |  |
| Minimal | 4 patients | 1 patient |  |
| Moderate | 2 patients | 3 patients |  |
| Large | 0 patients | 0 patients |  |
| **Treatment Factors** |  |  |  |
| Type of chemotherapy regimens*** |  |  | 0.936 * |
| FOLFOX | 5 patients | 9 patients |  |
| Gemcitabine / (nab)-paclitaxel | 4 patients | 5 patients |  |
| FOLFIRINOX | 4 patients | 4 patients |  |
| Other | 9 patients | 11 patients |  |
| Mean number of chemotherapy cycles per patient | 11 +/- 5 | 12 +/- 9 | 0.433 ** |
| Number of chemotherapy lines |  |  | 0.103 * |
| First-line | 9 patients | 4 patients |  |
| Second-line | 5 patients | 12 patients |  |
| Third-line | 3 patients | 2 patients |  |
| Timing of chemotherapy |  |  | 1.000 * |
| Perioperative | 5 patients | 6 patients |  |
| Postoperative only | 9 patients | 10 patients |  |
| Preoperative only | 3 patients | 2 patients |  |

\* two-tailed Fisher's exact test, \*\* two-tailed Student's t-test, \*\*\* more than one regimen per patient possible

### Formation of Study Groups Based on Chemotherapy Response

The only significant predictor of cancer-specific survival within the study cohort was the number of chemotherapy cycles administered (p=0.009, Figure 1). Cancer type (p=0.281), the extent of the cancer measured by the peritoneal cancer index (p=0.335), and the patients’ performance status measured by ECOG score (p=0.203) did not correlate with survival. Because of these findings, three strata were created based on the total number of chemotherapy cycles administered (1-6 cycles (n=7), 7-12 cycles (n=19), >12 cycles (n=9)). Patients were assigned to study groups depending on whether their actual cancer-specific survival was greater than the median survival within their stratum (i.e., sensitive, n=17) or less (i.e., resistant, n=18). All patients who were alive at the last follow-up had a follow-up greater than the median survival within its stratum. There were no significant differences in patient, disease, and treatment factors between study groups (Table 1). Of the 1010 image patches from the dataset, 500 were labeled sensitive and 510 resistant.

**Figure 1:**
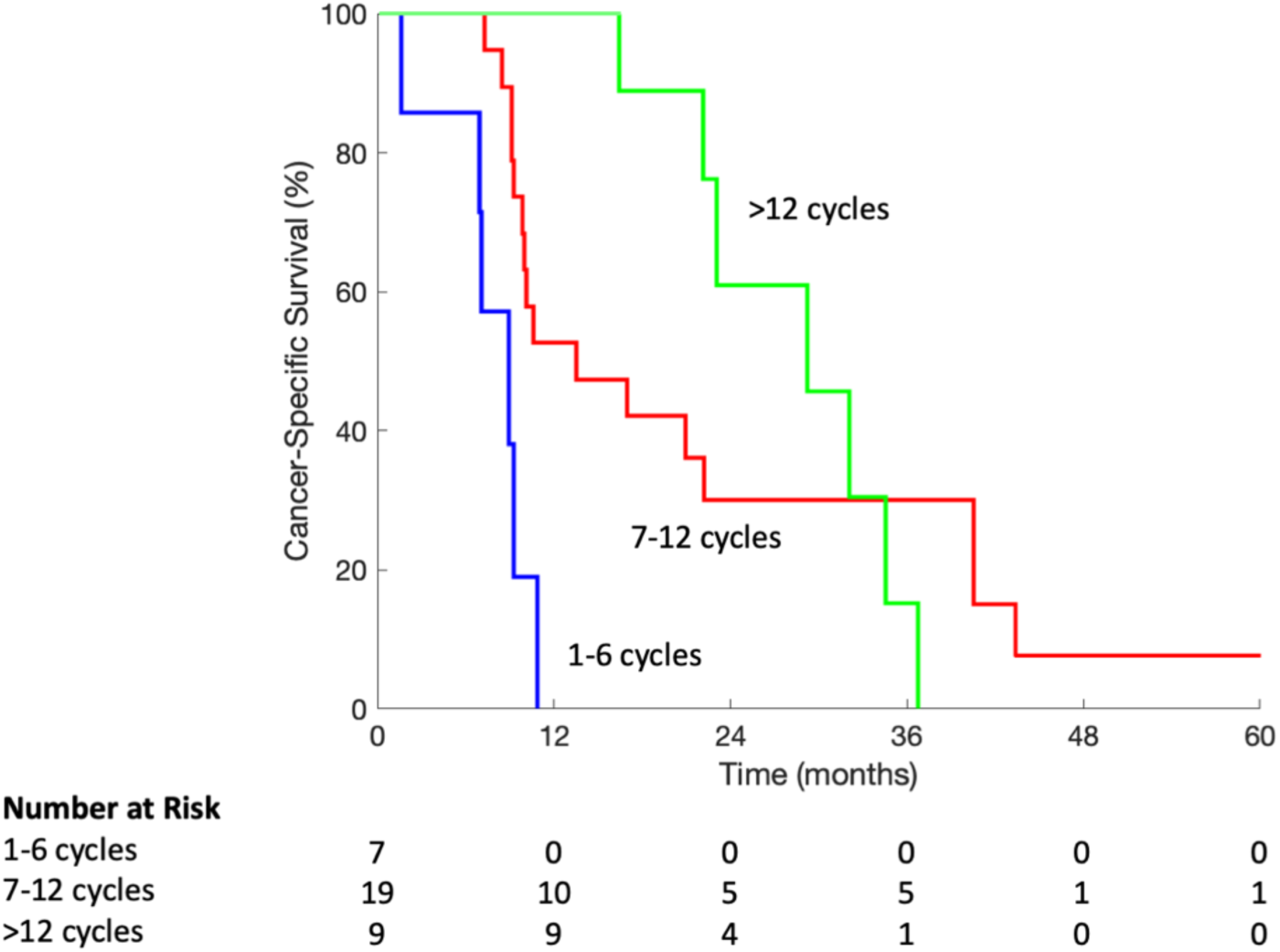
Cancer-specific survival stratified according to the total number of chemotherapy cycles administered.

### System Development and Its Architecture

Initially, we tested a wide range of well-established convolutional neural networks (CNNs) typically used for image classification through supervised learning without image augmentation or any other modification (simple CNN, DarkNet, EfficientNet, GoogLeNet, SqueezeNet, ResNet-50). The systems were trained and evaluated using a random 80:20 train-test split on a lesion level. With a prediction threshold of 0.5, the accuracy of these networks in correctly predicting the presence or absence of chemotherapy resistance to standard chemotherapy regimens was less than 0.54.

The poor performance of these off-the-shelf systems was likely due to the relatively small number of images within the dataset (1010 images from 101 biopsied lesions). To address these issues, we subsequently developed a densely connected CNN based on Huang at al.^18^ and provided modifications with relatively fewer layers. The dataset was split into training and testing sets at random with an 80:20 ratio assigned on a patient level. Preprocessing of the training set included augmentation of the images to add more variability for better generalization^19^. The network architecture included three densely connected convolutional blocks (output from all previous layers was input to subsequent layers within the same block) with transition convolution layers between the blocks. We added dropout layers that helped to prevent overfitting during training^20^. 10-fold cross-validation^21^ was applied. For each test image patch, the system provided a predicted probability in the range of 0.0 to 1.0 of whether an image patch belonged to a patient with chemotherapy resistance. For performance assessment, a prediction threshold of 0.5 was used. Under this scheme, we trained 10 models of an identical densely connected CNN architecture, with the performance reported as the mean across all 10 folds.

### System Performance on Images of Biopsy-Confirmed Metastases

The mean accuracy of the developed deep learning system in correctly predicting chemotherapy resistance on individual PSM image patches was 0.79 (95%CI 0.77-0.82, sensitivity 0.70, specificity 0.89, AUC-ROC 0.77, Figure 2). Since each PSM’s weight of contribution to the patient’s outcome cannot be predicted, we used a majority voting scheme to compute a patient label from all the PSM image patches within a patient. Under this approach, the accuracy of predicting chemotherapy resistance of individual patients was 0.80 (95%CI 0.63-0.92, sensitivity 0.72, specificity 0.88, AUC-ROC 0.78, Figure 2).

**Figure 2:**
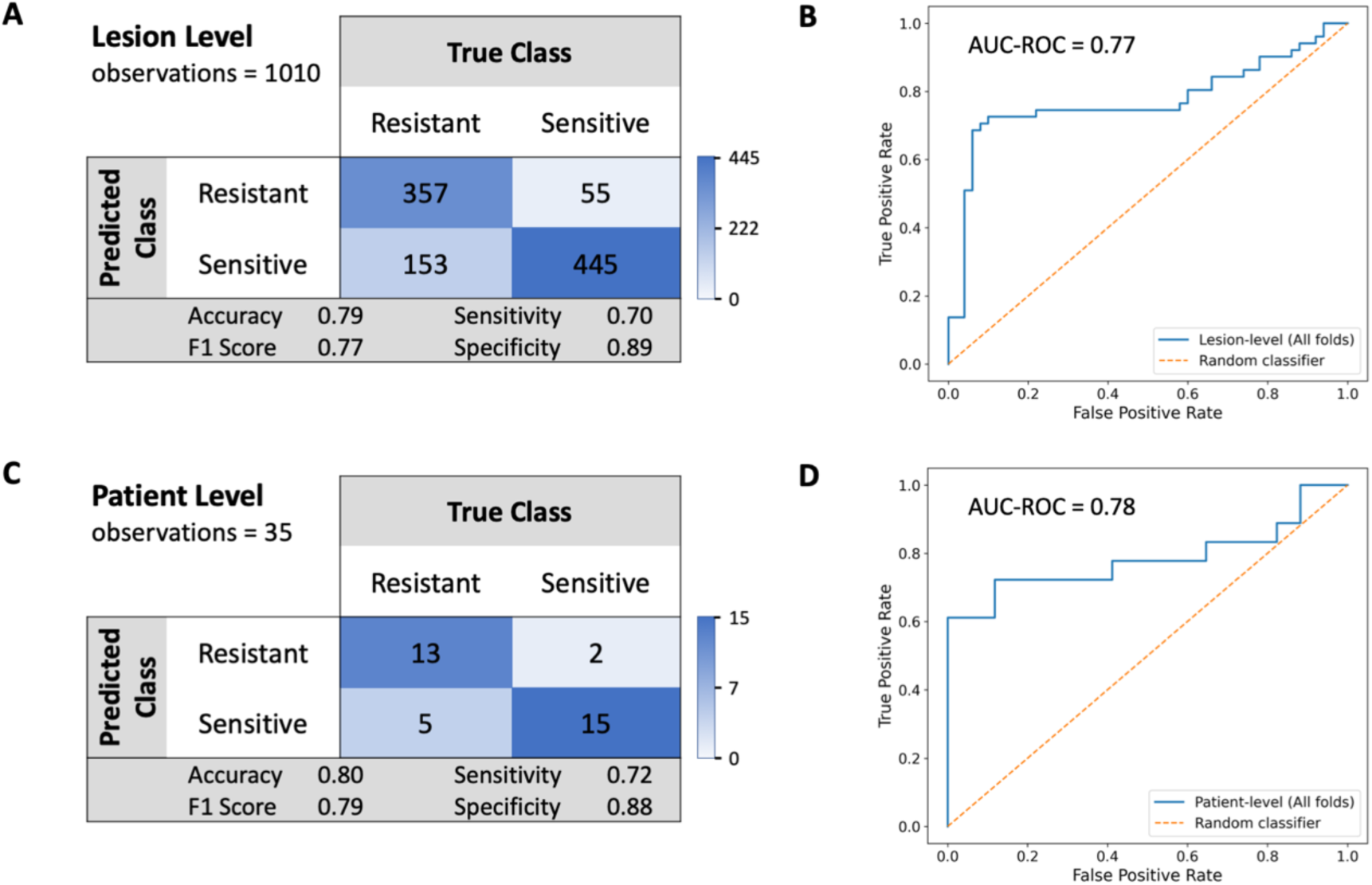
Confusion matrix of the densely connected CNN’s performance on 1010 image patches using 10-fold cross-validation predicting each PSM’s class (A). Receiver operating characteristic curve of predicted probability of a PSM belonged to a patient with chemotherapy resistance (B). Similar assessment of the system’s performance using a majority voting scheme to compute a patient label from all the PSM image patches within a patient (C, D).

### Saliency Maps for Trustworthiness Assessment

The 10-fold cross-validation produced ten models of which six achieved 100% accuracy in predicting patient labels, two achieved 75%, one achieved 50%, and one achieved 33%, respectively. To assess the system’s trustworthiness, Grad-CAM^22^ saliency maps were generated using the model whose accuracy was closest to the 10-fold mean and had the smallest train-test gap (Figure 3). These maps suggested the model identifies sensitive PSMs using features within the lesion itself (72% of relevant prediction pixels were inside the PSM), whereas it distinguishes resistant PSMs through features in the immediately surrounding area near blood vessels (73% of relevant prediction pixels were outside the PSM). The findings emphasized class-specific spatial feature utilization rather than random attention.

**Figure 3:**
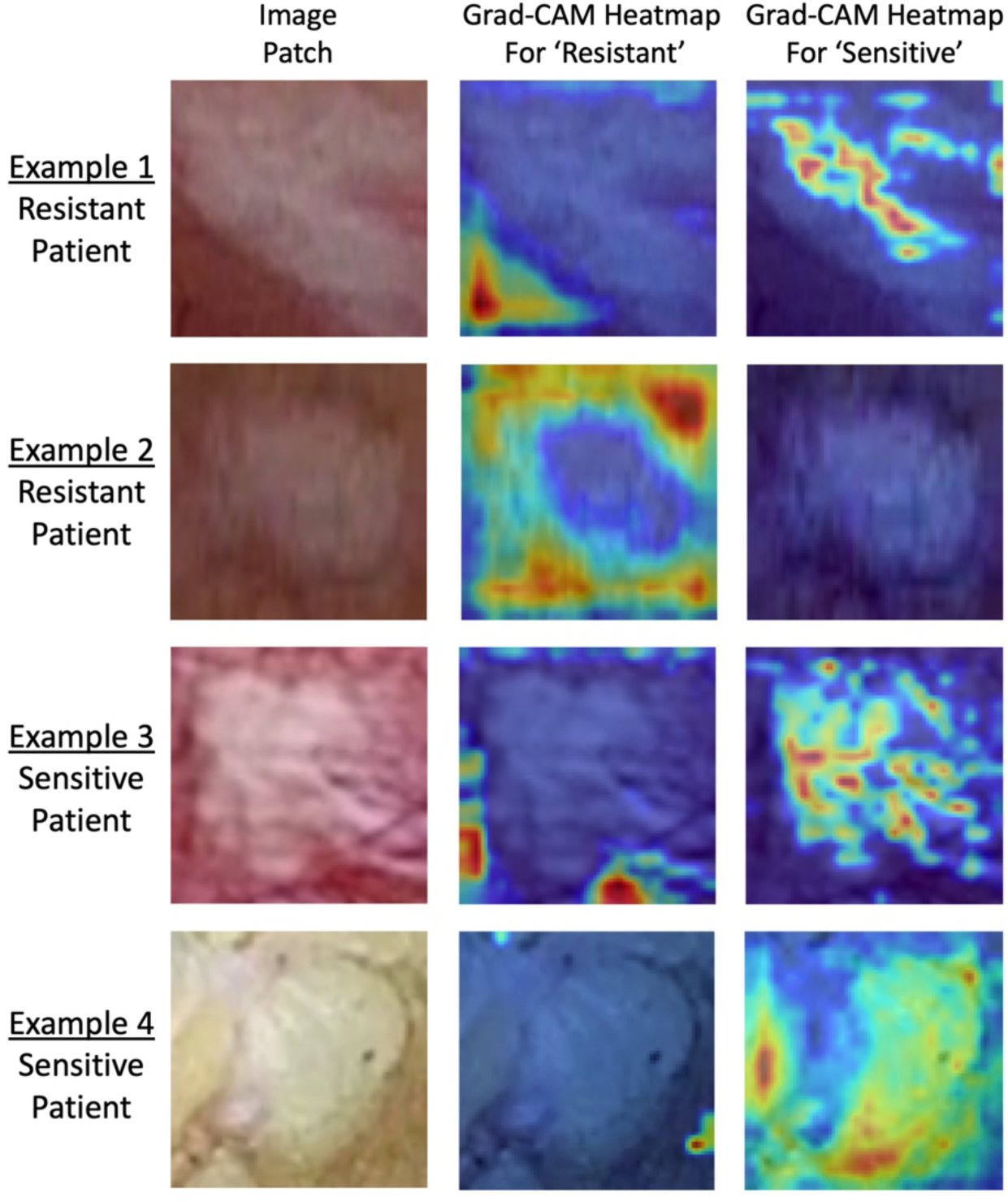
Examples of Grad-Cam saliency maps highlighting the regions that increase the score for a specific class.

## Discussion

This study demonstrated the technical feasibility of a novel approach of using standard laparoscopy images of PSMs in their natural environment to predict individualized chemotherapy responses through a densely connected CNN. Grad-CAM analysis suggested class-specific spatial attention patterns, with predictions for chemotherapy-sensitive and chemotherapy-resistant PSMs relying on distinct intratumoral and peritumoral regions, respectively, thereby supporting the interpretability and trustworthiness of the system’s decision-making process. It raised the hypothesis that chemotherapy resistance could be associated with changes in the structural appearance of small blood vessels supplying the cancer tissue in the peritumoral region; a feature that could only be assessed in its natural environment and, therefore, requiring the method described. The current system may have only learned some relevant features needed for generalizable performance and therefore will require additional training data with a wider distribution and variability. Yet, support for such an approach comes from Ma et al.^23^ who demonstrated in a feasibility study the potential to predict disease-free survival using laparoscopy images in patients with advanced ovarian cancer. While their study did not measure chemotherapy resistance (i.e., their model was not controlled for other factors that influence disease-free survival, such as extent of disease and type of treatment provided (e.g., extent of debulking and rate of chemotherapy administered)), used whole image prediction (i.e., images depicted suspected cancer without biopsy confirmation along with a lot of surrounding healthy peritoneal surfaces, introducing noise into the model), and its potential risk for overfitting (i.e., a complex model not adjusted to the limited data variability (larger number of images from each video with expected similarities)), it still demonstrated the potential of a transformer encoder to make somewhat related predictions.

Implementation of our method would be feasible given the broad availability of standard laparoscopy, only requiring minor updates of their processors for such a system to run in real time. Even though a large proportion of patients with advanced gastrointestinal cancers currently do not undergo laparoscopy as part of routine care, considering this for such cohorts in the future would be reasonable given its perceived benefit for dictating the most effective chemotherapy regimen while at the same time providing a diagnostic biopsy (omitting the need for other forms of biopsies) through a procedure with minimally greater risk and clinical recovery compared to currently performed invasive biopsies. The provided information from such a system could fundamentally change the approach to metastatic gastrointestinal cancers by allowing oncologists to personalize treatment plans, spare patients from ineffective and toxic drugs, and prevent critical delays in administering more effective therapies.

Despite the promising results and the demonstrated theoretic feasibility of developing such a system, given the current stage of an early prototype in this retrospective observational feasibility study, there are several limitations. 1) Overfitting is a potential concern given that the study presents data from a relatively small sample size, images derived from a single surgeon’s experience, and the complex architecture of the system. To mitigate overfitting and improve generalization, the number of model layers was minimized, data augmentation was applied to increase training data variability, dropout layers and L2 regularization were incorporated, batch normalization was employed to stabilize training, and a learning rate scheduler was used to enhance convergence. 2) The sample is somewhat heterogenous with similar but different cancer types and similar but different types of chemotherapy administered, which provides some limitations in defining chemotherapy resistance. However, given that the demographics were similar between groups (Table 1), the impact of this limitation is likely small. 3) Different metastases within the same patient may respond differently to chemotherapy. At its current state, it is impossible to determine which metastases within a patient are dictating the patient’s survival. Hence, it is conceivable that in patients who responded poorly to chemotherapy, some of the PSM evaluated were indeed chemotherapy sensitive and vice versa. 4) Even with a system that can reliably predict chemotherapy resistance to standard chemotherapy regimens, such a system (at least at this stage) would not provide answers about specific drugs and would not be able to name the most appropriate alternative treatment. I.e., a patient labeled chemotherapy resistant is not guaranteed to do better with a different chemotherapy regimen. Naming the most effective regimen in any individual patient will need to be established through future iterations of the system. 5) The system at its current stage only applies to patients with PSMs from non-colon gastrointestinal adenocarcinoma and is not applicable to patients with other cancers or patients with isolated metastases in the depth of the liver or in the lung. Similarly, the study did not include patients with very advanced metastatic disease who usually do not undergo routine laparoscopy. Many of the above limitations are anticipated to be addressed in planned follow-up studies. In particular, multi-institutional studies evaluating each cancer type separately, undergoing one type of chemotherapy regimen in a large sample could help validate the results.

In conclusion, knowledge about chemotherapy resistance in individual patients with metastatic cancer is important to guide the choice of chemotherapy and the choice of alternative approaches. But even from a surgical perspective, it is important since it can guide the threshold of when to consider operative resection or debulking in the setting of metastatic disease that is expected to respond favorably to chemotherapy. Our feasibility study demonstrates a pathway of how to evaluate intra-operatively imaged PSMs and comes with tremendous technical promise. The future clinical space for such technology is broad and is not only limited to cancer patients who undergo routine laparoscopic operations. Depending on the performance of a more matured system, it could potentially be utilized for all patients with a clinical or radiographic high probability of PSMs. Yet, future applications will depend on results from validation studies. The current system still requires further development in a larger multi-institutional sample and subsequent external validation.

## Data Availability

Due to study patient confidentiality and privacy concerns, any data is only available upon written request. According to a standard access procedure, applications will be reviewed by our research administration for scientific merit, evaluated for the fit of the data for the proposed methodology, assessed for any conflict with current studies, and verified that the proposed use meets Good Clinical Practice guidelines. Investigators wishing to use any of our patient data are asked to submit a brief description of the proposed project by contacting. Investigators can expect an initial response within 6 weeks of request submission. Data would only be provided in deidentified format, after approval of the appropriate IRB, and after execution of a data use agreement. Our source code is available upon request by contacting.

## Author Contributions

Drs Schnelldorfer and Gaikwad had full access to all of the data in the study and take responsibility for the integrity of the data and the accuracy of the data analysis.

*Concept and design*: Schnelldorfer, Gaikwad.

*Acquisition, analysis, or interpretation of data*: All authors.

*Drafting of the manuscript*: Schnelldorfer.

*Critical review of the manuscript for important intellectual content*: All authors.

*Statistical analysis*: Schnelldorfer, Gaikwad.

*Obtained funding*: Schnelldorfer.

*Administrative, technical, or material support*: Schnelldorfer, Gaikwad.

*Supervision*: Schnelldorfer.

## Additional Contributions

We thank Georgios Georgalis, PhD (Tufts Institute for Artificial Intelligence), the study participants, operating room staff, and research administration who contributed to the study.

## References

1. Hu Z, Li Z, Ma Z, Curtis C. Multi-cancer analysis of clonality and the timing of systemic spread in paired primary tumors and metastases. Nat Genet 2020;52:701–708. doi:10.1038/s41588-020-0628-z

2. Guo H, Zhang J, Qin C, et al. Biomarker-targeted therapies in non-small cell lung cancer: current status and perspectives. Cells 2022;11:3200.

3. Kim JY, Jeon E, Kwon S, et al. Prediction of pathologic complete response to neoadjuvant chemotherapy using machine learning models in patients with breast cancer. Breast Cancer Res Treat 2021;189:747–757.

4. LeSavage BL, Suhar RA, Broguiere N, et al. Next-generation cancer organoids. Nat Mater 2022;21:143–159.

5. Shimizu H, Nakayama KI. Artificial intelligence in oncology. Cancer Sci 2020;111:1452–1460.

6. Tran KA, Kondrashova O, Bradley A, et al. Deep learning in cancer diagnosis, prognosis and treatment selection. Genome Med 2021;13:152.

7. Bhinder B, Gilvary C, Madhukar NS, Elemento O. Artificial intelligence in cancer research and precision medicine. Cancer Discov 2021;11:900–915.

8. Maaref A, Romero FP, Montagnon E, et al. Predicting the response to FOLFOX-based chemotherapy regimen from untreated liver metastases on baseline CT: a deep neural network approach. J Digit Imaging 2020;33:937–945.

9. Wei J, Cheng J, Gu D, et al. Deep learning-based radiomics predicts response to chemotherapy in colorectal liver metastases. Med Phys 2021;48:513–522.

10. Nakanishi R, Oki E, Hasuda H, et al. Radiomics texture analysis for the identification of colorectal liver metastases sensitive to first-line oxaliplatin-based chemotherapy. Ann Surg Oncol 2021;28:2975–2985.

11. Li C, Qin Y, Zhang WH, et al. Deep learning-based AI model for signet-ring cell carcinoma diagnosis and chemotherapy response prediction in gastric cancer. Med Phys 2022;49:1535–1546.

12. Hayashi K, Ono Y, Takamatsu M, et al. Prediction of recurrence pattern of pancreatic cancer post-pancreatic surgery using histology-based supervised machine learning algorithms: a single-center retrospective study. Ann Surg Oncol 2022;29:4624–4634.

13. Bai M, Liu M, Wang S, Ran M, Ye J, Xie Y, Zhang C, Wang Y, Lu J, Dai F, Li D, Li H. Organoids in cancer therapy: translational applications and clinical promise. Mol Cancer 2026. doi: 10.1186/s12943-026-02678-7.

14. Heckelmann B, Duhn J, Braun R. Current advances in preclinical patient-derived cultivation models for individualized drug response prediction in pancreatic cancer. Oncol Res 2026;34:1. doi: 10.32604/or.2026.075028.

15. Tsimberidou AM, Fountzilas E, Nikanjam M, Kurzrock R. Review of precision cancer medicine: Evolution of the treatment paradigm. Cancer Treat Rev 2020;86:102019.

16. Fountzilas E, Tsimberidou AM, Vo HH, Kurzrock R. Clinical trial design in the era of precision medicine. Genome Med 2022;14:101.

17. Schnelldorfer T, Castro J, Goldar-Najafi A, Liu L. Development of a deep learning system for intra-operative identification of cancer metastases. Ann Surg 2024; 280:1006–1013. doi:10.1097/SLA.0000000000006294

18. Huang G, Liu Z, Van Der Maaten L, Weinberger KQ. Densely connected convolutional networks. Proceedings of the IEEE Conference on Computer Vision and Pattern Recognition 2017:2261–2269. doi:10.1109/CVPR.2017.243

19. Mikołajczyk A, Grochowski M. Data augmentation for improving deep learning in image classification problem. IEEE International Interdisciplinary PhD Workshop 2018:117–122.

20. Srivastava N, Hinton G, Krizhevsky A, et al. Dropout: a simple way to prevent neural networks from overfitting. J Mach Learn Res 2014;15:1929–1958. doi:10.5555/2627435.2670313

21. Wong TT. Performance evaluation of classification algorithms by k-fold and leave-one-out cross validation. Pattern Recognition 2015;48:2839–2846. doi:10.1016/j.patcog.2015.03.009

22. Selvaraju RR, Cogswell M, Das A, Vedantam R, Parikh D, Batra D. Grad-CAM: Visual explanations from deep networks via gradient-based localization. Proc IEEE Int Conf Comput Vis 2017:618–626. doi:10.1109/ICCV.2017.74

23. Ma X, Hsu YC, Asare A, Zhang K, Glassman D, Handley KF, Foster K, Sharma K, Westin S, Jazaeri A, Fleming ND, Bhattacharya PK, Jiang X, Sood AK, Shams S. A pioneering artificial intelligence tool to predict treatment outcomes in ovarian cancer via diagnostic laparoscopy. Sci Rep 2025;15:14437. doi:10.1038/s41598-025-98434-w

